# Procrastination and Test Anxiety among Medical Students in Medical Colleges in Nepal: A Cross-Sectional Study

**DOI:** 10.1101/2025.08.01.25332587

**Authors:** Rashmi Kandel, Sujan Babu Marahatta, Sudip Khanal, Bimala Poudel, K.C Sadikshya

**Author notes:** **Corresponding Author:** Rashmi Kandel, or. Rashmi Kandel and Sujan Babu Marahatta contributed equally to this work.

## Abstract

Procrastination among student’s is a prevalent behavior that negatively influences their performance and well-being. This research assessed with the prevalence of medical students struggle with their behavioral trait of procrastinating and test anxiety in Nepalese students.

A cross-sectional study was conducted among 214 medical students (MBBS) of Chitwan Medical College through pretested semi-structured questionnaire through self-administered questionnaire. The questionnaire was self-developed with added The Academic Procrastination Scale and The Westside Test Anxiety Scale aiming assess the psychological and behavioral trait of medical students. The collected data were entered and analyzed using SPSS IBM v.20. Data were analyzed using chi-square test, t-test, ANOVA, logistic regression and correlation where association were established where p value was <0.05. All ethical consideration were followed and IRC was approved with registration: **NEHCO/IRC/080/051, MMIHS**.

Prevalence of Test Anxiety (TA) was found to be 39.3% among studied population. TA among medical students has strong association with maternal and paternal education level with significance. In this study, respondents who had maternal and paternal education level secondary/more (95% CI 0.526-2.056, p-value=0.036 and 95% CI 0.463-2.476, p-value=0.048) respectively are more likely to have TA. The factors such as residence, alcohol use and smoking and/or tobacco products/chewing were associated with the academic procrastination. The mean academic procrastination was significantly higher to those living close to campus on their own/with flat mates (p = 0.001), with individuals who consume alcohol displaying higher mean scores than those who do not (p = 0.008), individuals who use smoking and/or tobacco products/chewing have higher mean scores compared to non-users (p = 0.033).

The strong correlation between the academic procrastination and test anxiety with the positive correlation value (r=0.593) which illustrate that students with test anxiety is greatly affected by the academic procrastination. Regular assessments and counseling services offered by colleges can effectively manage among students.

## Introduction

### Background

Procrastination is defined as ‘to voluntarily delay an intended course of action despite expecting to be worse off for the delay’.(1) Procrastination is a pervasive phenomenon in academic contexts with a range of negative outcomes.(2),(3) Procrastination among student’s is a prevalent behavior that negatively influences their performance and well-being.(4)

Test Anxiety is an intense psychological state experienced by test-takers concerning the evaluation of their test performance and possible consequences that would happen in their personal or academic lives after test results.(5) The phenomenon of test anxiety (TA) is composed of two distinct elements: cognitive components (such as worry and test-irrelevant thinking) and affective components (such as emotionality and bodily symptoms).(6) The cognitive aspect of worry refers to the negative thoughts test-takers have regarding the potential of failing a test and its consequences, the affective aspect of emotionality encompasses physiological responses (such as headache and increased heartbeat) as well as feelings of nervousness and tension. Failure in this particular situation encompasses not solely the uncertainty about securing the minimum required grade or pass mark but also the incapacity to meet the requirements for their career progression or to fulfill the expectations of one’s parents, among other factors.(7)

Surveys were carried out as part of the International College Student project were the World Mental Health International College Student (WMH-ICS) Initiative acknowledged that the college years holds great significance as it marks the transition from the late-adolescent stage to adulthood for students. It is notable that this transition occurs during a highly delicate phase of the life cycle, when the prevalence of emotional difficulties and mental illnesses commonly occur. Epidemiological investigations reveal that a considerable proportion, ranging from 25% to 40%, of university students experience test anxiety.(8),(9) Various research has shown that people who procrastinate are not as healthy as people who do not procrastinate.(10)

Studies based on Canada and US suggest high prevalence of depression, anxiety and psychological distress among medical students than in general population.(11) Test anxiety is considered as a significant issue among medical students, as it has high probability of leading to underachievement, low performance, demotivation, and psychological distress.(12) Procrastination shows a correlation with negative behaviors and consequences, such as late submission of assignments, cramming, anxiety regarding examinations and social interactions, use of self-handicapping strategies, fear of failure, underachievement, and the potential damage in mental well-being, specifically depression and anxiety.(13)

### Statement of Problems

Procrastination was most prevalent among students in higher education, with estimates that college students engage in this behavior 60% of the time.(14) Among undergraduate students, epidemiologic studies shows that 25 to 40% experience test anxiety.(8),(9)

### Global Scenario

Evidence further indicates that medical school programs are characterized by challenging classes and high credit loads in each academic semester, thereby correlating with higher levels of test anxiety, resulting poor academic performance and increased rates of school dropout.(15) In Saudi Arabia the study conducted where the occurrence of test anxiety among students before examinations was at a significantly high rate of 53.04%. Similarly, psychological distress was prevalent among students at a rate of 82.50%. Furthermore, there was a noteworthy positive linear correlation (p<0.01) between test anxiety and psychological distress as the years of the course progress.(16)

A cross-sectional correlational investigation was conducted on 317 medical students at Shiraz University of Medical Sciences; Iran showed the findings that 29.25% of the students had academic procrastination (nearly always or always). Furthermore, a substantial 47.9% of the students reported that engaging in academic procrastination at a moderate level resulted a lot of difficulties for them.(17) About 65% students experienced exam anxiety due to various reasons major one which is medical course.(18) Since the medical course being so vast and examination process so lengthy, students’ perception of course load and their ability to manage time with their course work was associated with exam anxiety.(18)

Univariate analyses demonstrated that procrastination exhibited a correlation with low mindfulness, high stress, and poor perceived health. To investigate the connections between procrastination and stress, as well as procrastination and perceived health, the researchers employed structural equation modelling to that revealed that the effects were mediated by mindfulness, and bootstrapping analyses confirmed the significance of these effects.(19) A survey conducted among 220 nursing students at a large regional university of USA exhibit moderate test anxiety where 100 individuals were classified as the low test anxiety group, while 120 individuals were classified as the moderate-high cognitive test anxiety group. As a result, 55% of the nursing students who participated in the survey demonstrated symptoms of cognitive test anxiety.(20) A study conducted in Nyeri District, Kenya prevalence was found to be 68.1%.(21)

### Situation in South East Asia

The study among medical students who were undertaking their Ophthalmology clinical examination at Penang Medical College, test anxiety is a significant concern as it can affect students’ performance and well-being during examinations.(12) Research findings of Malaysia showed, 22.2% of pharmacy students categorized of “moderately high” test anxiety while only 7.6% of them have “high” level of test anxiety. However, findings suggested that moderately high test anxiety rate (32.6%) and high prevalence rate of psychological distress (61.1%) existed among pharmacy students.(22)

The study found a statistically significant, positive correlation between test anxiety score and procrastination score among physiotherapy students, mean procrastination score was 74.1.(23) The cross-correlations conducted among Chinese students, between academic procrastination and test anxiety exhibited positive and significant associations at T1 (r =.314, p <.01) and T2 (r =.175, p <.01). Moreover, the T1 academic procrastination demonstrated a positive correlation with T2 test anxiety (r =.477, p <.01), while the correlation between T1 test anxiety and T2 academic procrastination was relatively weak (r =.165, p <.05). In order to ascertain whether T2 test anxiety and T2 academic procrastination could be predicted independently by T1 academic procrastination and T1 test anxiety, a comprehensive cross-lagged panel analysis was undertaken.(24)

The research conducted in India revealed that the prevalence of high exam anxiety among Phases I-III were 37%, 28%, and 32% respectively. It was observed that males exhibited higher levels of exam anxiety compared to females. The examination structure, inadequate time management, and the extensive curriculum were identified as the primary factors contributing to anxiety associated with the examination.(25)

### Situation in Nepal

In the examination of the study, factors played an important role in psychological morbidity, where the crucial psychosocial factors included the ‘quality of food in mess’, ‘high parental expectations’, ‘lack of entertainment’, ‘feeling of loneliness’, and ‘worrying about the future’ and these factors may be linked to staying in the hostel.(26) Prevalence of psychological morbidity was 20.9% and was higher among students of basic sciences, Indian nationality and whose parents were medical doctors.(26)

Those pursuing a medical degree faced challenges with encompassing academic demands, psychological strains, and existential pressures.(27) In context of Nepalese study, it was found high prevalence of poor mental health among medical students of Nepal and future studies required to identify the factors associated with poor mental health.(28)

## Methods

### Study design

The study design for the research on “Procrastination and Test Anxiety among Medical students in Medical Colleges” was cross-sectional study. The study assessed the prevalence of Procrastination and Test Anxiety among medical students along with finding out association among Procrastination and Test Anxiety. Also, the study found association between dependent and independent variables.

### Study Method

Quantitative method was conducted where data collected through self-administered questionnaire was analyzed using statistical, mathematical or numerical methods. Findings were visualized using charts and tables.

### Study time

The study was conducted from Ashad to Chaitra 2080 B.S. During the course of these ten months, all the activities including formulation of research problem, proposal development, data collection and analysis and also preparation of report for the dissemination of findings was done.

### Study Site

The study site for the research was selected to be Chitwan. Medical college of Chitwan hosts a significant number of medical students who are likely to experience test anxiety and procrastination due to the rigorous nature of their academic program which allowed generalization of findings among medical students in national context.

### Study Population

The study population of this research were students enrolled in medical colleges. Students from each year first to fifth were included studying MBBS in medical colleges of Chitwan excluding year third due to situational circumstances.

### Study unit

The study unit were individuals who were students studying MBBS in CMC.

### Sample size

To determine the sample size, following formula was used:

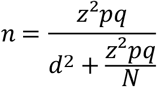

Where, n = Desired sample size

Z = Standard normal deviate, usually set at 1.96 which corresponds to 95% confidence level

p = Sample proportion

=50%

=0.5

q = 1-p

=50%

=0.5

N = Total population

d = Allowable error

=5% (0.05)

So, 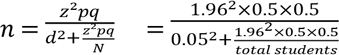

Again,

Taking 10% as non-response rate, sample size becomes,

### Sampling Technique

Multistage proportionate sampling was the method used for sampling purpose in the study. First of all, the study area medical colleges of Chitwan were purposively selected. A complete list of all students of medical colleges was obtained from college. There were total students (of all year’s 1^st^, 2^nd^, 4^th^ and 5^th^) in the sampling frame in this study.

**Figure 1:**
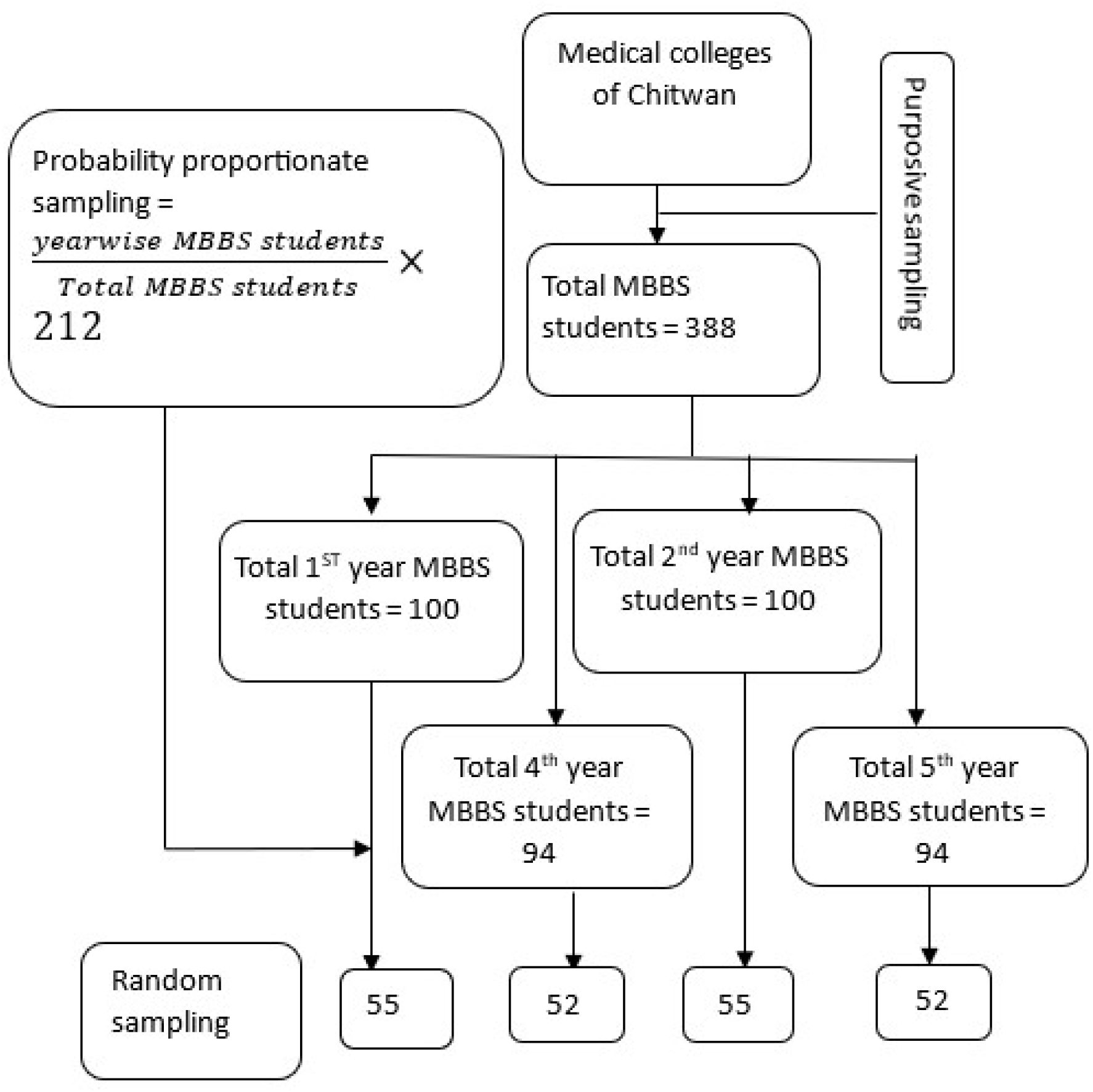
Sampling Strategy

## Criteria for sample collection

Based on the following criteria, the sample were selected:

### Inclusion Criteria

MBBS students of first, second, fourth and fifth year enrolled in medical colleges of Chitwan and excluding year third due to situational circumstances.

### Exclusion Criteria

Students who do not voluntarily agree to participate were excluded from the study. Student absent on the date of data collection were excluded from the study.

### Data collection tools

Semi-structured questionnaire was used to collect quantitative data from MBBS students to assess the prevalence of procrastination and test anxiety among medical students. The questionnaire was adopted based on The Academic Procrastination Scale and The Westside Test Anxiety Scale. The questionnaire also included sociodemographic information and individual factors.

### Data collection techniques

The semi-structured questionnaire was distributed among MBBS students where the students were given to self-administered the questions.

### Pretesting the data collection tools

Pretesting of the questionnaire was done among 24 MBBS students (10% of the sample population) enrolled in Nepal Medical College.

### Validity and Reliability of the study tools

#### Validity

For the validity of the study, proposal preparation and validated tools was drafted in close guidance of the supervisor. Questionnaire were adopted from the validated tools. Also, the questionnaire was developed based on various previously conducted studies. Adequate literature was reviewed throughout the study to ensure external validity of the study. The set of questionnaires was checked and verified by concerned teachers.

#### Reliability

For the reliability of the study tools, pretesting of the tools was done in similar population at non study area. The questionnaire was modified and corrected based on feedback. A total of 10% of the filled questionnaire were cross-checked to test the reliability of the study.

### Ethical Consideration

Ethical approval was obtained from Institutional Review Committee (IRC), MMIHS. Similarly, permission and approval from the Chitwan Medical College administration was obtained prior to the data collection. Consent form was filled by the students prior to the data collection which accumulates the information about research. The participants were given the liberty to voluntarily participate in the study and withdraw from it any time. The participants were informed and oriented about the purposes of the study. Study participants were assured about the confidentiality and non-disclosure of information for purposes other than research.

### Data management and analysis

The collected data was organized, coded, and entered in SPSS and MS-excel. In this study The Academic Procrastination Scale which contained 25 items. Each item was measured on a scale of 1 to 5, where response of 1 indicates strongly disagreement and 5 as strongly agreement to the statement. In the questionnaire items 1,8,12,14 and 25 indicated reverse-scored items. Here, independent t-test or ANOVA, were applied to compare the mean scores of different variables. The Westside Test Anxiety Scale which contained 10 items. Each item was measured on scale of 1 to 5, where response of 1 indicates not at all and 5 as extreme agreement to the statement. Here, Chi-square test along with binary and multiple logistic regression were applied. Correlation among procrastination and test anxiety was also explored and p-value less than 0.005 was considered significant.

### Dissemination of research results

Research presentation in college and submission of research report to Institutional Review Committee (IRC). Research findings disseminated to the concerned authorities and stakeholders. The research approached to be published in Journal.

## Results

A cross-sectional descriptive and analytical study with multistage proportionate sampling was carried out in 214 participants to assess procrastination and test anxiety among medical students in medical colleges was conducted in Chitwan Medical College, Bharatpur Chitwan. The prevalence of procrastination and test anxiety along with their respective associated factors were identified by this study. The findings of the study were analyzed using various parametric test and SPSS software which is explained in this chapter.

### Socio-demographic characteristics

Table 3 illustrates the sociodemographic characteristics of the study participants. The age distribution of the respondents, with a median age of 21.50 and an interquartile range of 3, the age range of 18 to 26 years. Among 214 participants, more than two third of the respondents (63.6%) were male in comparison to female respondents (36.4%). Regarding ethnicity, the majority identified as more than two fifth Brahmin (42.5%) and roughly a quarter Madhesi (29.0%), with smaller proportions representing Chhetri (11.2%), Janjati (10.7%), Muslim (4.2%), and Dalit (2.3%) ethnic groups. The almost all of the respondents held Nepali nationality (93.0%), with a smaller fraction being Indian (7.0%).

**Table 1:** Age, Sex, Ethnicity and Nationality related characteristics.

| **Sociodemographic characteristics** | **Number (n=214)** | **Percentage (%)** |
| --- | --- | --- |
| **Age (In completed years)** |  |  |
| **(Median ± IQR= 21.50 ± 3)** |  |  |
| 18-20 | 65 | 30.4 |
| 21-26 | 149 | 69.6 |
| **Sex** |  |  |
| Male | 136 | 63.6 |
| Female | 78 | 36.4 |
| **Ethnicity** |  |  |
| Brahmin | 91 | 42.5 |
| Chhetri | 24 | 11.2 |
| Janjati | 23 | 10.7 |
| Dalit | 5 | 2.3 |
| Madhesi | 62 | 29.0 |
| Muslim | 9 | 4.2 |
| **Nationality** |  |  |
| Indian | 15 | 7.0 |
| Nepali | 199 | 93.0 |

**Table 2:**
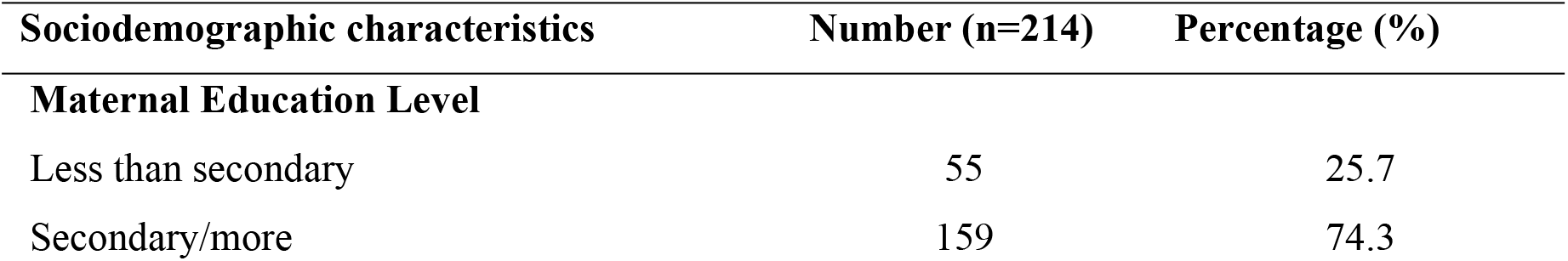
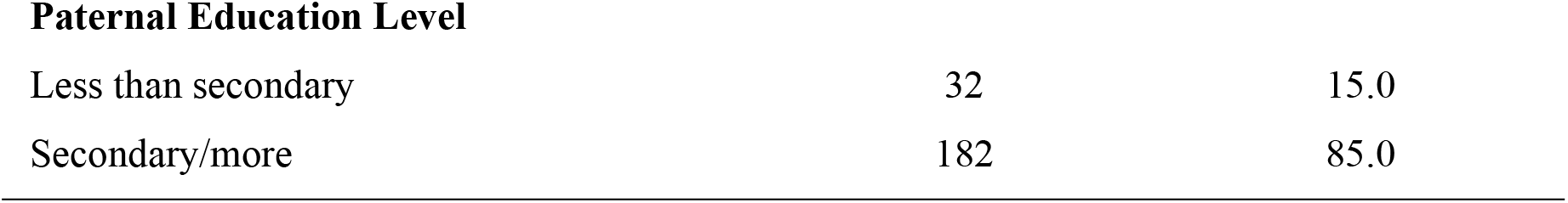
Parental Educational Level related characteristics.

| **Sociodemographic characteristics** | **Number (n=214)** | **Percentage (%)** |
| --- | --- | --- |
| **Maternal Education Level** |  |  |
| Less than secondary | 55 | 25.7 |
| Secondary/more | 159 | 74.3 |
| **Paternal Education Level** |  |  |
| Less than secondary | 32 | 15.0 |
| Secondary/more | 182 | 85.0 |

**Table 3:**
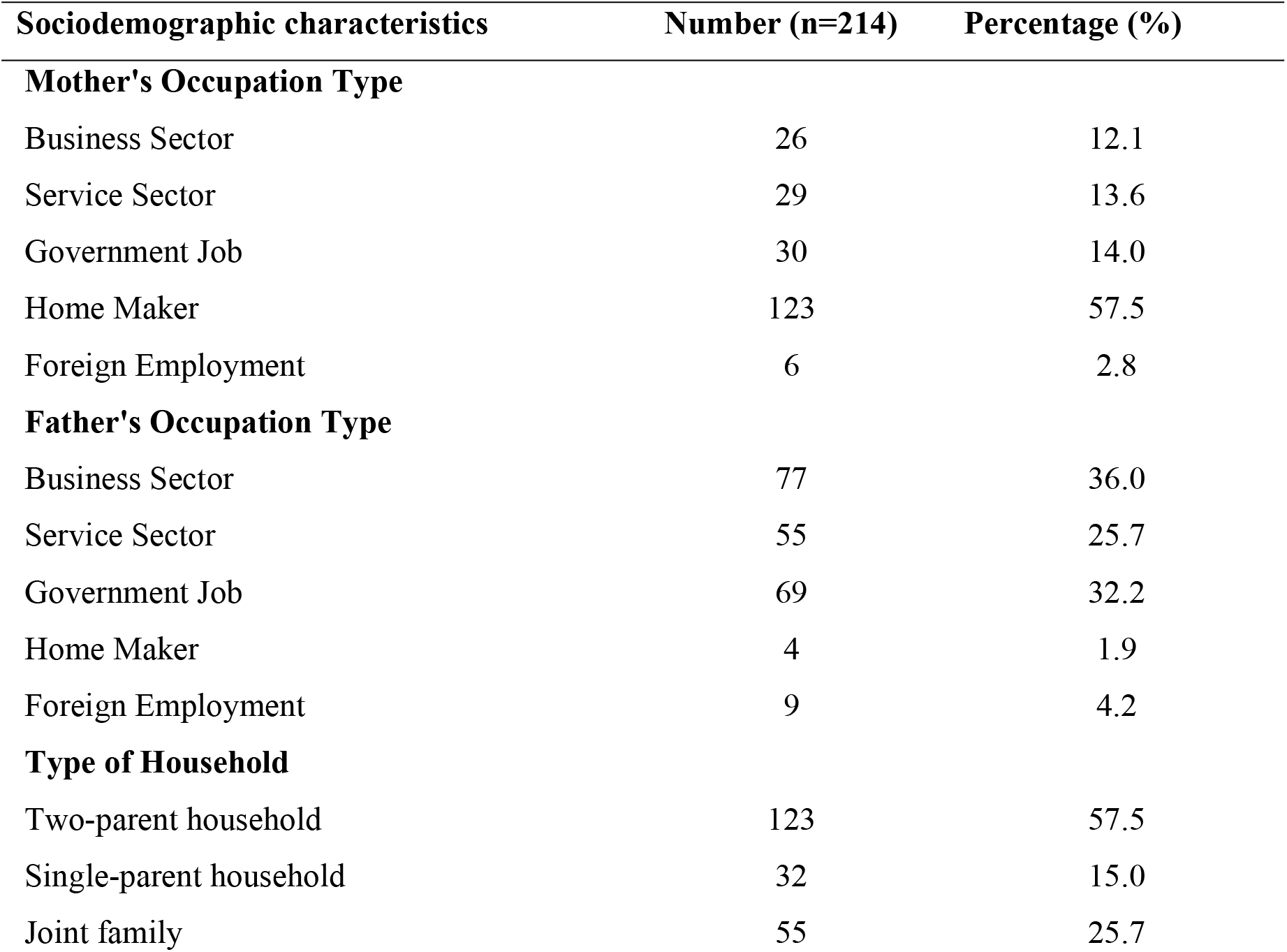
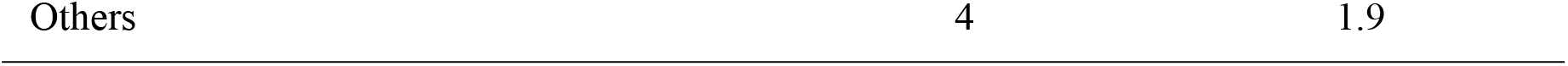
Types of Parental Occupation and Household.

**Table 4:** Individual factors of respondents.

| Characteristics | Number (n=214) | Percentage (%) |
| --- | --- | --- |
| **Year of Education (currently in)** |  |  |
| First year | 54 | 25.2 |
| Second year | 56 | 26.2 |
| Fourth year | 52 | 24.3 |
| Fifth year | 52 | 24.3 |
| **Residence (currently in)** |  |  |
| Hostel | 106 | 49.5 |
| Lives close to campus on their own/with flat mates | 75 | 35.0 |
| Lives at home with family | 33 | 15.4 |
| **Alcohol Use** |  |  |
| Yes | 21 | 9.8 |
| No | 193 | 90.2 |
| **Smoking and/or Tobacco Product (chewing) Use** |  |  |
| Yes | 11 | 5.1 |
| No | 203 | 94.9 |

**Table 5:**
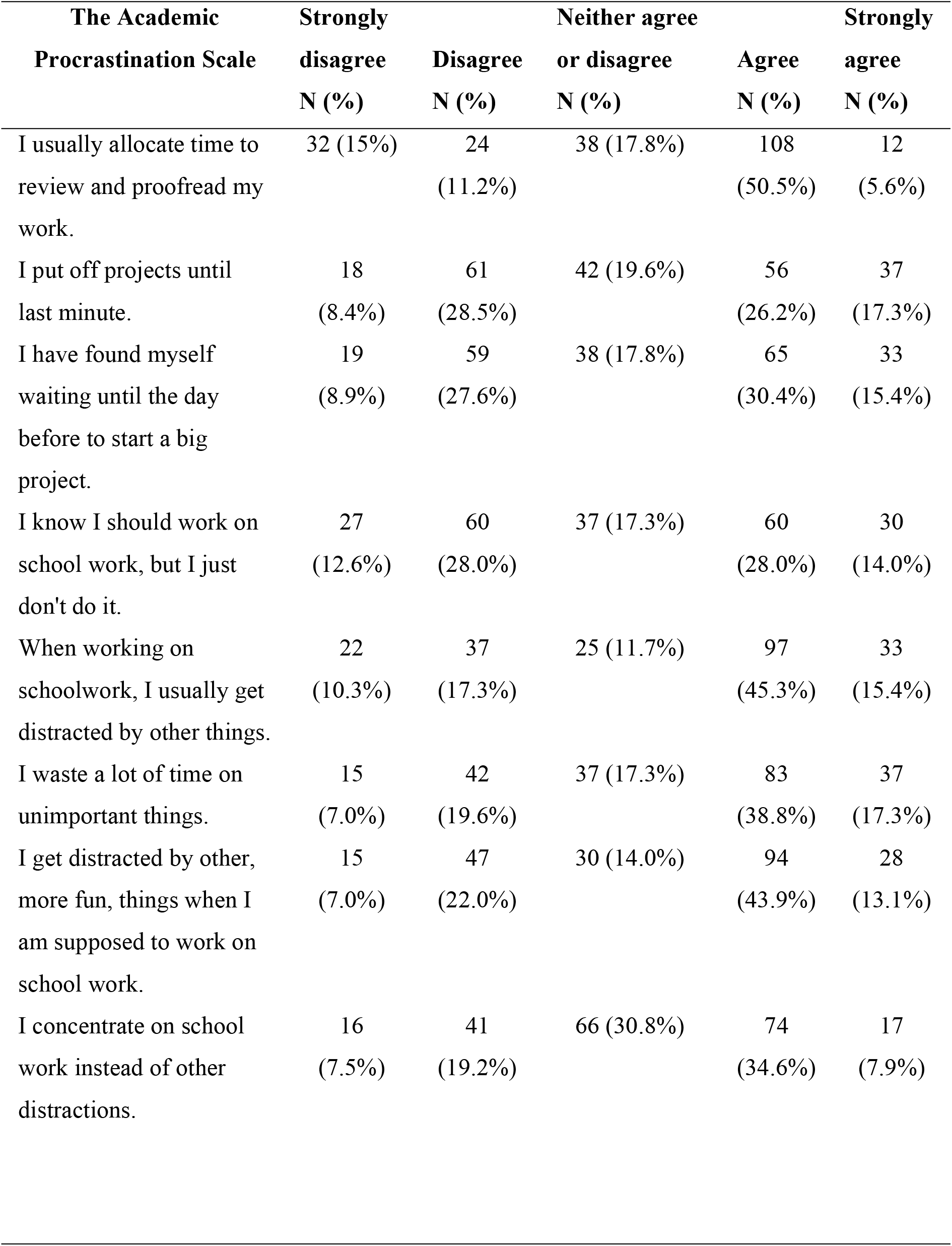
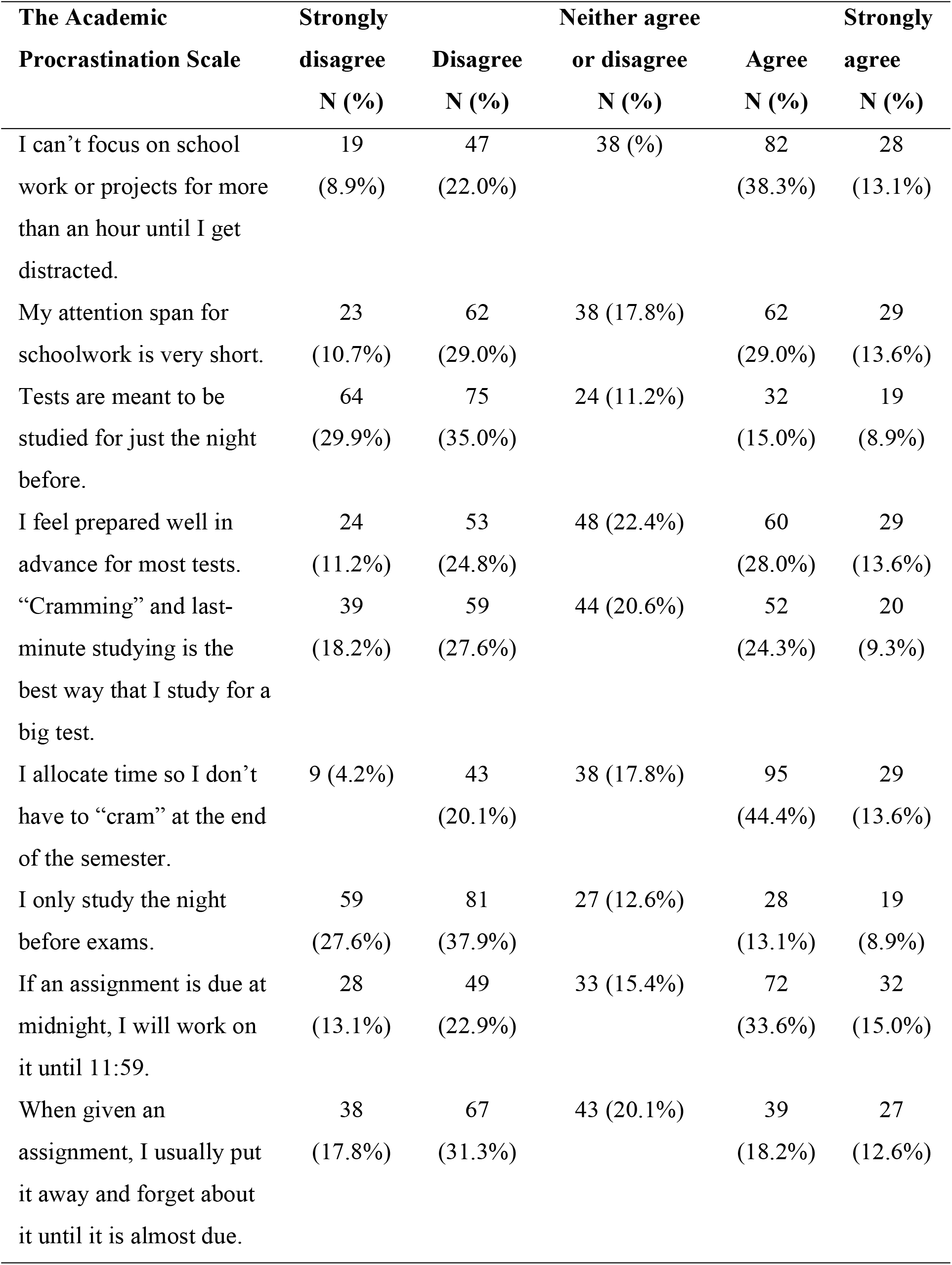
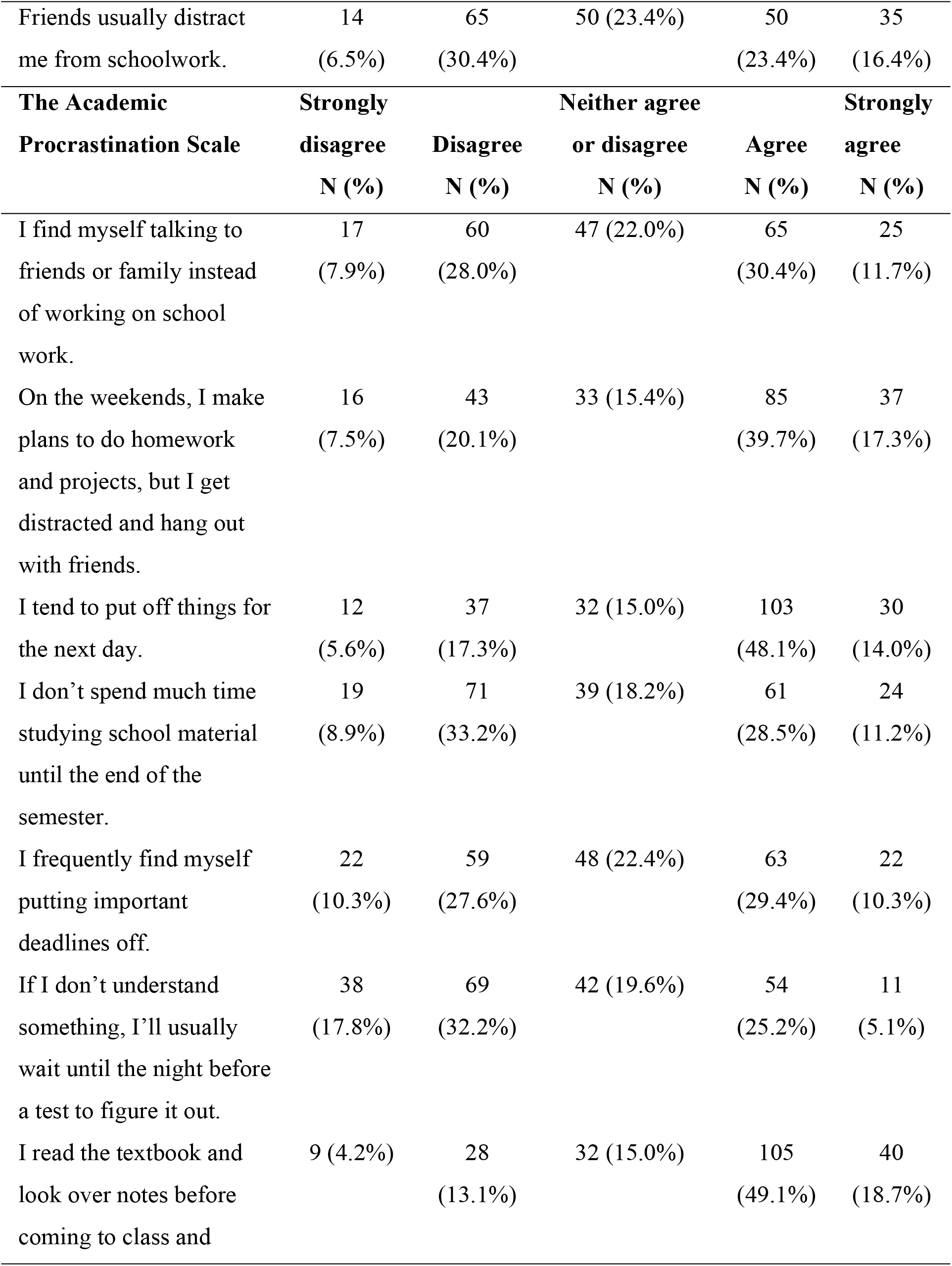
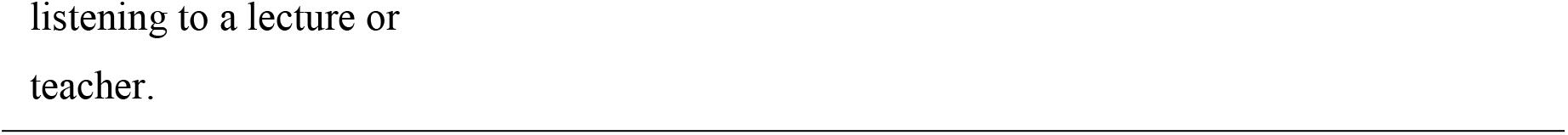
The Academic Procrastination Scale.

**Table 6:**
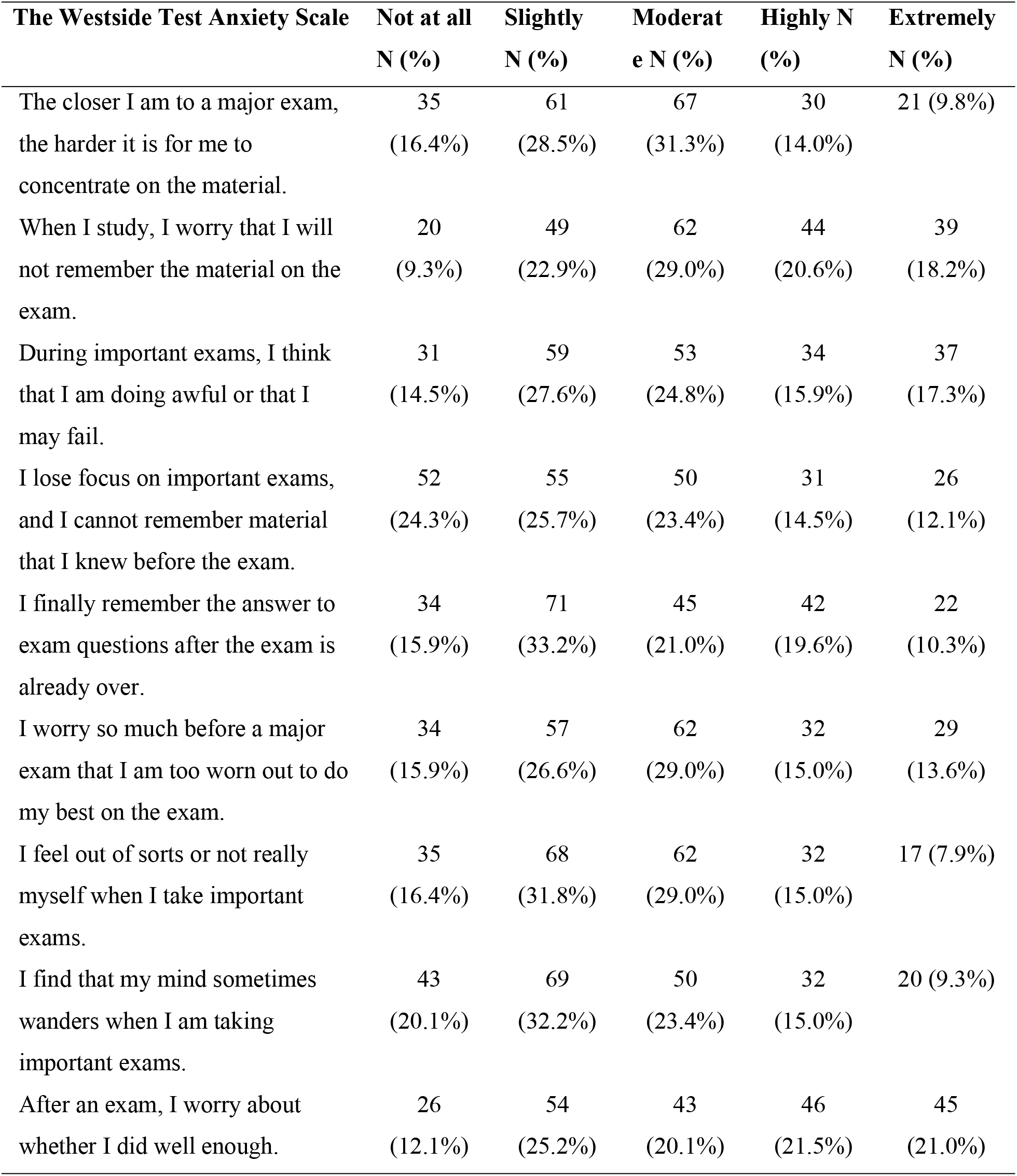
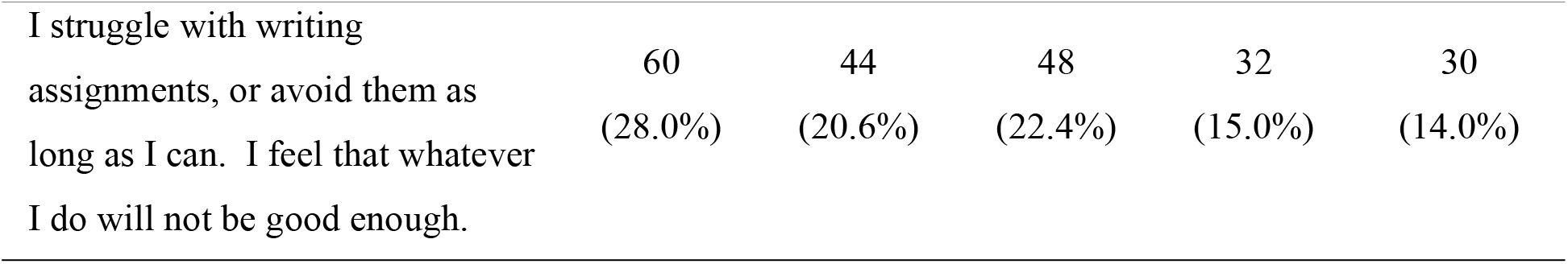
The Westside Test Anxiety Scale.

**Table 7:** Association between Academic Procrastination and Sociodemographic characteristics.

| Characteristics | Mean | SD | SE | t/f | p-value |
| --- | --- | --- | --- | --- | --- |
| **Age** |  |  |  |  |  |
| 18-20 | 70.46 | 16.066 | 1.993 | -0.710 | 0.478 |
| 21-26 | 71.97 | 13.498 | 1.106 |  |  |
| **Sex** |  |  |  |  |  |
| Male | 72.35 | 13.949 | 1.196 | 1.134 | 0.258 |
| Female | 70.05 | 14.884 | 1.685 |  |  |
| **Ethnicity** |  |  |  |  |  |
| Brahmin | 72.22 | 14.197 | 1.488 |  |  |
| Chhetri | 70.46 | 13.981 | 2.854 | 0.72 | 0.579 |
| Janjati | 68.78 | 15.406 | 3.212 |  |  |
| Madhesi | 70.82 | 13.900 | 1.765 |  |  |
| Others | 76.29 | 16.060 | 4.292 |  |  |
| **Nationality** |  |  |  |  |  |
| Nepali | 71.56 | 14.111 | 1.000 | 0.163 | 0.871 |
| Indian | 70.93 | 17.227 | 4.448 |  |  |

**Table 8:** Association between Academic Procrastination and Parental Education Level.

| Characteristics | Mean | SD | SE | t/f | p-value |
| --- | --- | --- | --- | --- | --- |
| **Maternal Education Level** |  |  |  |  |  |
| Less than secondary | 70.05 | 16.368 | 2.207 | -0.877 | 0.381 |
| Secondary/more | 72.02 | 13.539 | 1.074 |  |  |
| **Paternal Education Level** |  |  |  |  |  |
| Less than secondary | 68.78 | 15.024 | 2.656 | -1.173 | 0.242 |
| Secondary/more | 71.99 | 14.163 | 1.050 |  |  |

**Table 9:** Association between Academic Procrastination and Types of Parental Occupation and Household.

| Characteristics | Mean | SD | SE | t/f | p-value |
| --- | --- | --- | --- | --- | --- |
| **Mother's Occupation Type** |  |  |  |  |  |
| Business Sector | 69.96 | 15.534 | 3.047 | 0.392 | 0.814 |
| Service Sector | 71.97 | 14.529 | 2.698 |  |  |
| Government Job | 73.00 | 14.434 | 2.635 |  |  |
| Home Maker | 71.11 | 13.888 | 1.252 |  |  |
| Foreign Employment | 76.83 | 18.681 | 7.626 |  |  |
| **Father's Occupation Type** |  |  |  |  |  |
| Business Sector | 73.25 | 15.144 | 1.726 | 0.663 | 0.619 |
| Service Sector | 71.93 | 17.464 | 3.361 |  |  |
| Government Job | 69.65 | 12.133 | 1.461 |  |  |
| Home Maker | 70.50 | 22.098 | 11.049 |  |  |
| Foreign Employment | 68.89 | 13.148 | 4.383 |  |  |
| **Type of Household** |  |  |  |  |  |
| Two-parent household | 69.86 | 13.914 | 1.255 | 2.018 | 0.135 |
| Joint family | 74.20 | 15.534 | 2.095 |  |  |
| Others | 73.06 | 13.221 | 2.204 |  |  |

**Table 10:**
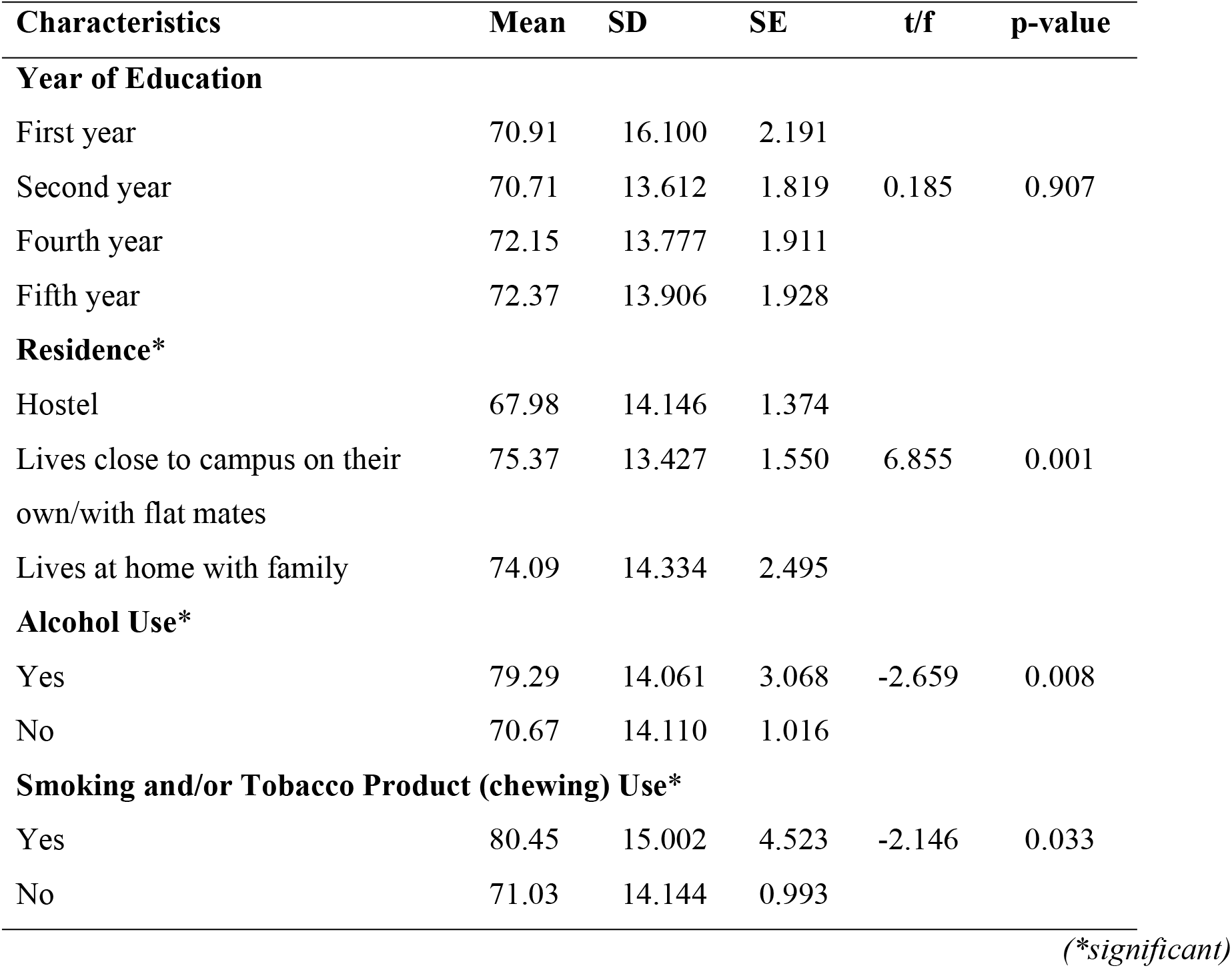
Association between Academic Procrastination and Individual Factor.

### Parental Educational Level related characteristics

Among the respondents, for maternal education level nearly three fourth (74.3%) had completed secondary education or higher, whereas 25.7% had less than secondary education. Similarly, for paternal education level, eight out of ten (85.0%) had attained secondary education or higher, while 15.0% had less than secondary education.

### Types of Parental Occupation and Household

Looking at occupation types among mothers, the more than half were homemakers, constituting (57.5%), followed by those in government jobs (14.0%), the service sector (13.6%), the business sector (12.1%), and foreign employment (2.8%). Conversely, among fathers, more than one third proportion was engaged in the business sector (36.0%), followed by government jobs (32.2%), the service sector (25.7%), and foreign employment (4.2%) whereas small percentage of fathers (1.9%) were homemakers. Household types varied, more than half with (57.5%) residing in two-parent households, 25.7% in joint families, 15.0% in single-parent households, and the remaining 1.9% in other arrangements such living with relatives and in hostel.

### Individual Factor of respondents

Table data provided highlights various individual factors among the respondents. Regarding the year of education, the distribution shows that one fourth (25.2%) are in their first year, followed closely by 26.2% in their second year, with similar proportions in the fourth and fifth years, each comprising 24.3%. In terms of residence, nearly half of the respondents (49.5%) live in hostels, while 35.0% reside close to campus either independently or with flat mates, and 15.4% live at home with their families. When it comes to alcohol use, the majority (90.2%) reported not consuming alcohol, while 9.8% indicated they did. Similarly, the majority (94.9%) reported no use of smoking or tobacco products, with only 5.1% acknowledging their use.

### The Academic Procrastination Scale

The Academic Procrastination Scale gives insights into the study habits and tendencies of the respondents. Notably, the more than half (50.5%) allocate time to review and proofread their work, indicating a proactive approach to academic tasks. However, more than one fourth (26.2%) admits to putting off projects until the last minute. Similarly, less than one third (30.4%) acknowledge waiting until the day before to start a big project. Despite recognizing the importance of schoolwork, more than one fourth (28.0%) confesses to not engaging in it as they should. Additionally, distractions during schoolwork are prevalent, with less than half (45.3%) reporting getting distracted by other things. Less than two fifth (38.8%) admits to wasting a considerable amount of time on unimportant tasks whereas, distractions from more enjoyable activities during study time are prevalent, with 43.9% reporting such occurrences. On the contrary, very few (7.9%) demonstrates a strong ability to concentrate on schoolwork despite distractions. However, less than two fifth (38.3%) struggles to maintain focus on academic tasks for extended periods, often getting distracted within an hour. Additionally, less than one third (29.9%) believe in studying for tests only the night before, suggesting a reliance on last-minute preparation methods. Despite this, more than one fourth (28.0%) express confidence in being well-prepared for exams in advance. Furthermore, while some (18.2%) resort to cramming and last-minute studying for important tests, others (13.6%) adopt more organized study approaches by allocating time for study so they don’t have to cram at the end. While more than one third (33.6%) admit to working on assignments right up to the deadline, indicating for last-minute completion, another less than one fifth (15.4%) tends to procrastinate by putting off assignments until they are nearly due. Moreover, social distractions seem to pose significant challenges, with more than one fifth (23.4%) reporting that friends typically divert their attention from schoolwork, and 30.4% finding themselves engaged in conversations with friends or family instead of focusing on academic tasks. Additionally, weekend plans often interfere with academic responsibilities for less than two fifth (39.7%), highlighting a tendency to prioritize social activities over homework and projects. Furthermore, procrastination extends to postponing tasks for the next day, with less than half (48.1%) admitting to this behavior. Similarly, more than one fourth (28.5%) acknowledges not dedicating enough time studying school material until the end of the semester, indicating a pattern of delayed academic engagement. Here, less than one third (29.4%) admits to frequently putting off important deadlines, while more than one fourth (25.2%) acknowledges waiting until the night before a test to address any areas of confusion. Interestingly, less than half (49.1%) report actively engaging with course material by reading the textbook and reviewing notes before attending class or listening to a lecture.

### The Westside Test Anxiety Scale

The Westside Test Anxiety Scale explain the anxiety levels experienced by respondents during academic evaluations. The data reveals varying degrees of test-related anxiety, with less than one third (31.3%) experiencing moderate levels of difficulty concentrating on material as major exams approach. Moreover, more than one fourth (29.0%) express concerns about their ability to recall material during exams, indicating a fear of forgetting important information. During important exams, more than one fourth (27.6%) worry about their performance, with some (15.9%) even fearing failure. Additionally, more than one third (33.2%) reports experiencing a lapse in memory during exams, indicating a significant impact of anxiety on cognitive functioning. For some, worries about exams can be overwhelming, with less than one third (29%) feeling too worn out to perform their best due to pre-exam stress. Additionally, less than one third (31.8%) feels out of sorts during important exams, indicating a high level of anxiety. Many (32.2%) find their minds wandering during exams, which could impact their focus and performance. Post-exam, concerns about performance persist for more than one fifth (21%), reflecting ongoing anxiety about results. Furthermore, more than one fourth (28%) struggles with writing assignments, fearing that their efforts won’t meet expectations.

### Inferential Analysis

#### ANOVA (f-test) or t-test of The Academic Procrastination Scale

##### Association between Academic Procrastination and Sociodemographic characteristics

The table below shows that among different age groups, individuals aged 18-20 have a mean score of 70.46, while those aged 21-26 have a mean of 71.97, although the difference is not statistically significant (p = 0.478). Regarding gender, males have a mean score of 72.35, slightly higher than females at 70.05, but the difference is not significant (p = 0.258). In terms of ethnicity, Dalit and Muslim combined have higher mean score of 76.29 whereas Brahmins have 72.22, Madhesi and Chhetri have similar value of 70.82 and 70.46 respectively and Janjati have lower mean of 68.78, with no significant difference observed (p = 0.579). Among Nepali and Indian nationalities, no significant difference is found in mean scores (p = 0.871).

##### Association between Academic Procrastination and Parental Education Level

Maternal education level shows no significant difference in mean scores between those with less than secondary education and secondary/more education (p = 0.381). Paternal education level similarly does not exhibit a significant difference in mean scores between those with less than secondary education has and secondary/more education (p = 0.242).

##### Association between Academic Procrastination and Types of Parental Occupation and Household

Mother’s and father’s occupation types also do not show significant differences in mean scores across different categories. Lastly, the type of household indicates a slightly higher mean score for individuals in a joint family setting compared to a two-parent household, although the difference is not significant (p = 0.135).

##### Association between Academic Procrastination and Individual Factor

The individual factors analysis explains on various factors and their correlation with mean scores, standard deviation (SD), standard error (SE), t-value/f-value, and p-value. Among different years of education, there is no significant difference in mean scores observed between first-year (70.91), second-year (70.71), fourth-year (72.15), and fifth-year (72.37) students (p = 0.907). However, a significant difference is found based on residence, with individuals living in hostels (67.98) exhibiting lower mean scores compared to those living close to campus on their own/with flat mates (75.37) or at home with family (74.09) (p = 0.001*). Moreover, significant differences are noted concerning alcohol use, with individuals who consume alcohol (79.29) displaying higher mean scores than those who do not (70.67) (p = 0.008*). Similarly, individuals who use smoking and/or tobacco products/chewing (80.45) have higher mean scores compared to non-users (71.03), with the difference being statistically significant (p = 0.033*).

#### Chi-square test of The Westside Test Anxiety Scale

##### Association between Test Anxiety and Sociodemographic characteristics

There shows no any significance with age of the students, sex, ethnicity, nationality in the table below.

**Table 11:**
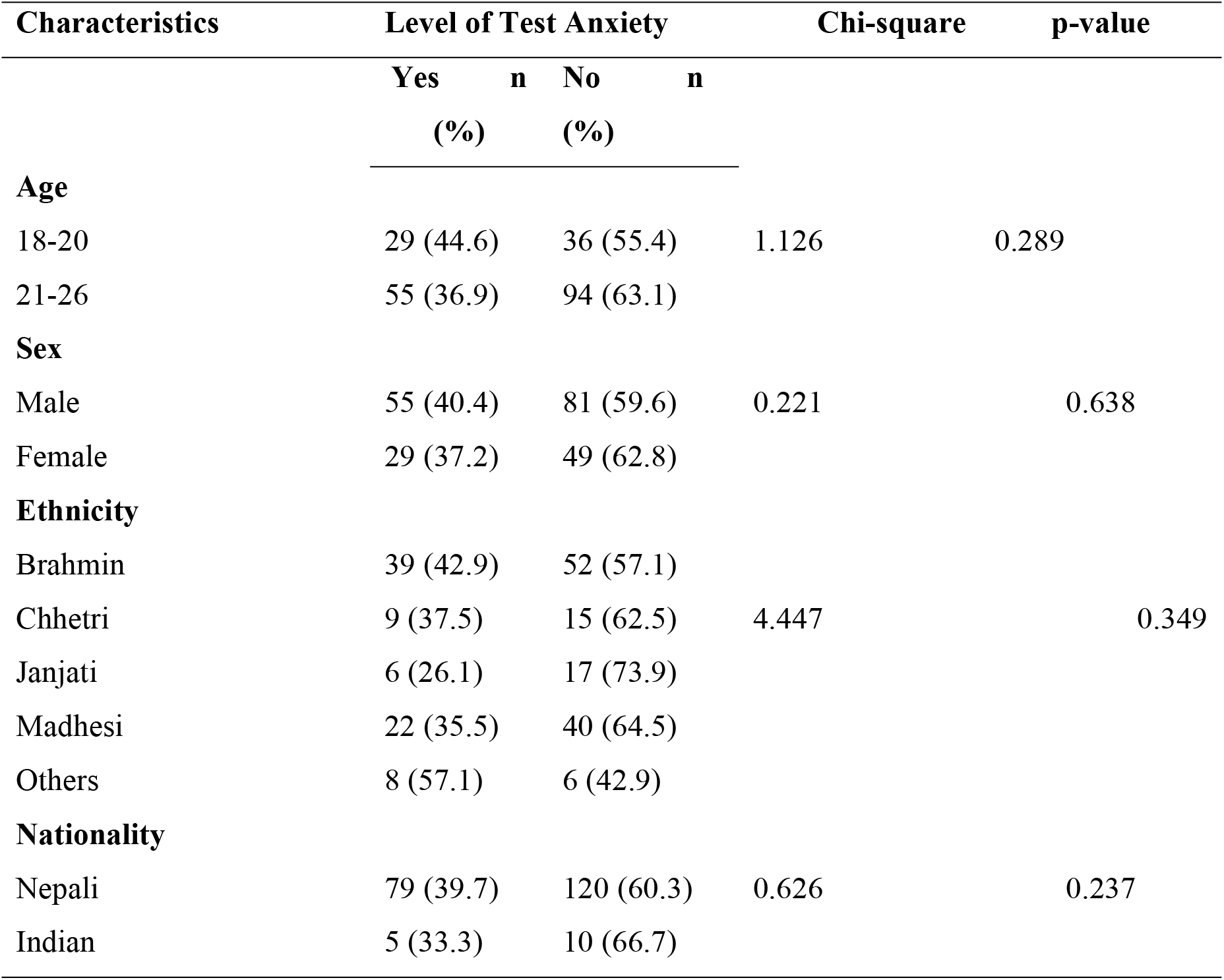
Association between Test Anxiety and Sociodemographic characteristics Characteristics Level of Test Anxiety Chi-square p-value.

##### Association between Test Anxiety and Parental Education Level

From the table 14 it reveals that Test Anxiety among medical students has strong association with maternal and paternal education level with a significance of p=0.036 and p=0.048 respectively.

**Table 12:** Association between Test Anxiety and Parental Education Level Characteristics Level of Test Anxiety Chi-square p-value.

| Characteristics | Level of Test Anxiety |  | Chi-square | p-value |
| --- | --- | --- | --- | --- |
|  | Yes<br>(%) | n No<br>(%) |  |  |
| **Maternal Education Level\*** |  |  |  |  |
| Less than secondary | 21 (38.2) | 34 (61.8) | 0.850 | 0.036 |
| Secondary/more | 63 (39.6) | 96 (60.4) |  |  |
| **Paternal Education Level\*** |  |  |  |  |
| Less than secondary | 12 (37.5) | 20 (62.5) | 0.826 | 0.048 |
| Secondary/more | 72 (39.6) | 110 (60.4) |  |  |

(\*significant)

**Table 13:**
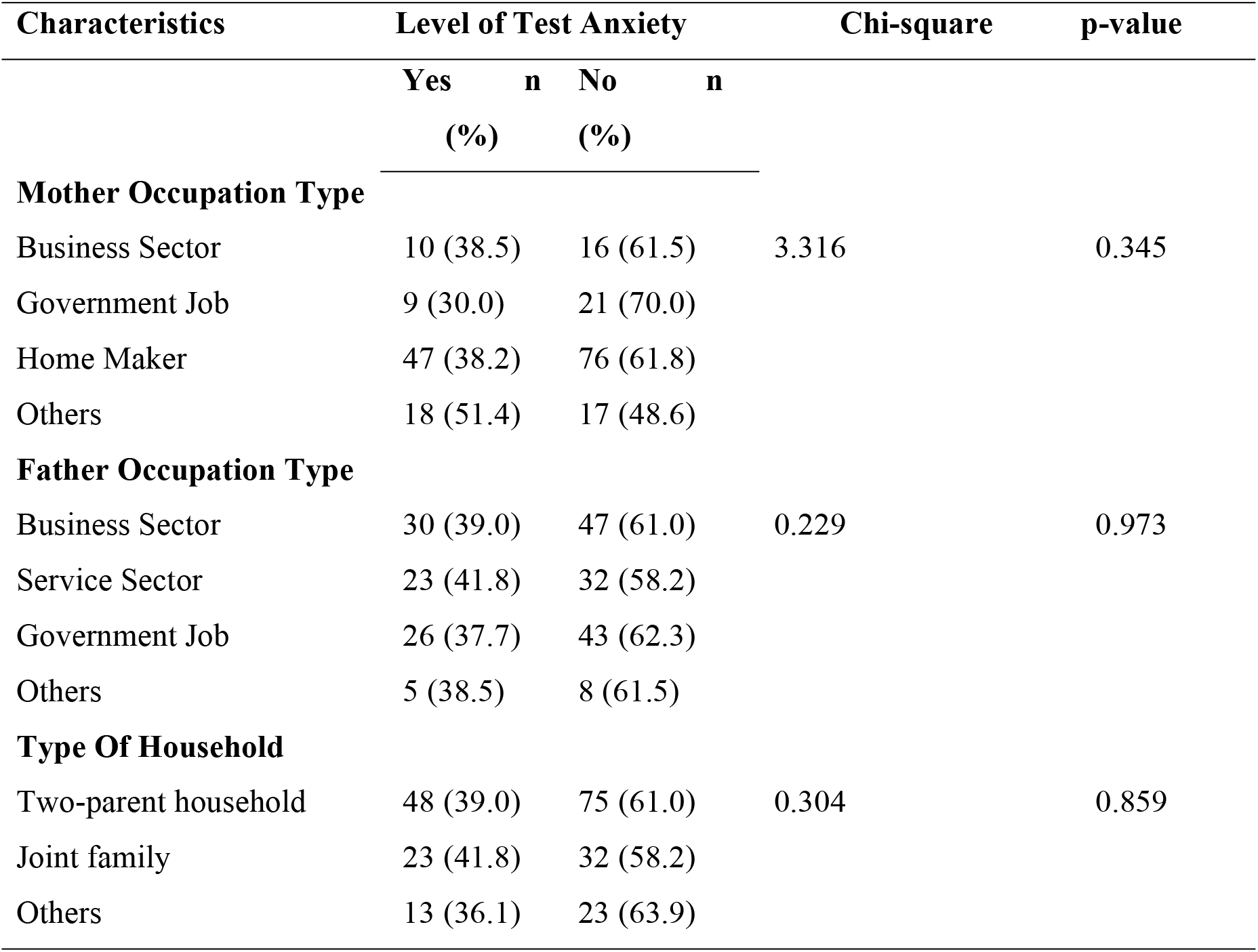
Association between Test Anxiety and Types of Parental Occupation and Household.

| Characteristics | Level of Test Anxiety |  | Chi-square | p-value |
| --- | --- | --- | --- | --- |
|  | Yes<br>(%) | No<br>(%) |  |  |
| **Mother Occupation Type** |  |  |  |  |
| Business Sector | 10 (38.5) | 16 (61.5) | 3.316 | 0.345 |
| Government Job | 9 (30.0) | 21 (70.0) |  |  |
| Home Maker | 47 (38.2) | 76 (61.8) |  |  |
| Others | 18 (51.4) | 17 (48.6) |  |  |
| **Father Occupation Type** |  |  |  |  |
| Business Sector | 30 (39.0) | 47 (61.0) | 0.229 | 0.973 |
| Service Sector | 23 (41.8) | 32 (58.2) |  |  |
| Government Job | 26 (37.7) | 43 (62.3) |  |  |
| Others | 5 (38.5) | 8 (61.5) |  |  |
| **Type Of Household** |  |  |  |  |
| Two-parent household | 48 (39.0) | 75 (61.0) | 0.304 | 0.859 |
| Joint family | 23 (41.8) | 32 (58.2) |  |  |
| Others | 13 (36.1) | 23 (63.9) |  |  |

**Table 14:**
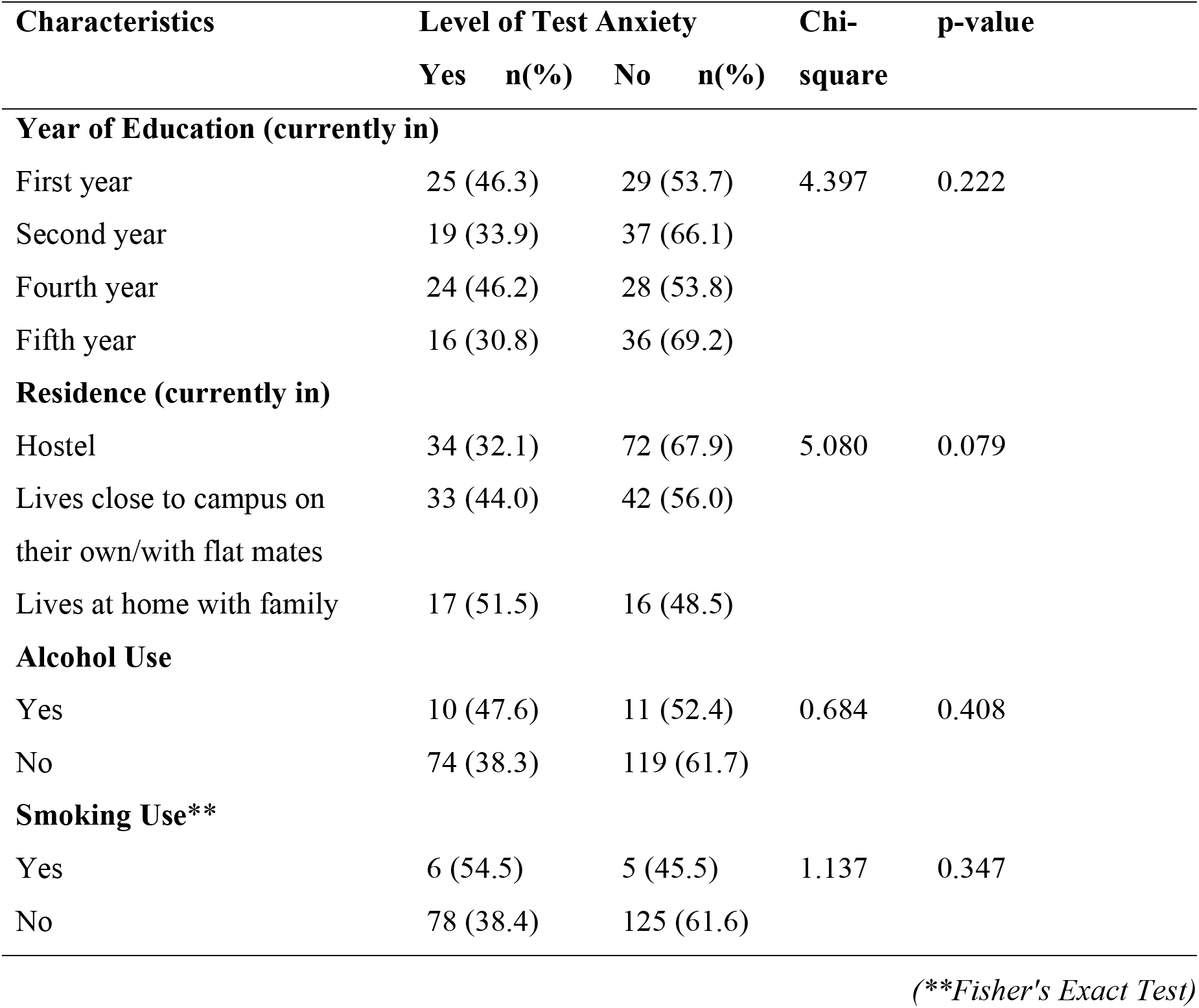
Association between Test Anxiety and Individual Factors.

##### Association between Test Anxiety and Types of Parental Occupation and Household

There shows no any association observed with mother, father occupation type and type of household as shown in table 15.

**Table 15:** Binary and Multiple logistic regression of Test Anxiety with Sociodemographic factors.

| Variable | Category | Test Anxiety |  | CO<br>R | 95%<br>CI | p-<br>valu<br>e | AO<br>R | 95%<br>CI | p-<br>valu<br>e |
| --- | --- | --- | --- | --- | --- | --- | --- | --- | --- |
| **Maternal**<br>**Educatio**<br>**n Level** | **Less than**<br>**secondary** | 21<br>(38.2<br>) | 34<br>(61.8<br>) | Ref |  | 0.85<br>0 | **Ref** |  | 0.91<br>0 |
|  | **Secondary/mor**<br>**e** | 63<br>(39.6<br>) | 93<br>(60.4<br>) | 1.06<br>2 | 0.566<br>-<br>1.995 |  | **1.04**<br>**0** | 0.526<br>-<br>2.056 |  |
| **Paternal**<br>**Educatio**<br>**n Level** | **Less than**<br>**secondary** | 12<br>(37.5<br>) | 20<br>(62.5<br>) | Ref |  | 0.82<br>6 | **Ref** |  | 0.87<br>3 |
|  | **Secondary/mor**<br>**e** | 72<br>(39.6<br>) | 110<br>(60.4<br>) | 1.09<br>1 | 0.503<br>-<br>2.368 |  | **1.07**<br>**1** | 0.463<br>-<br>2.476 |  |

##### Association between Test Anxiety and Individual Factors

From the table 16 it reveals that Test Anxiety among medical students has no any association with individual factors.

**Table 16:**
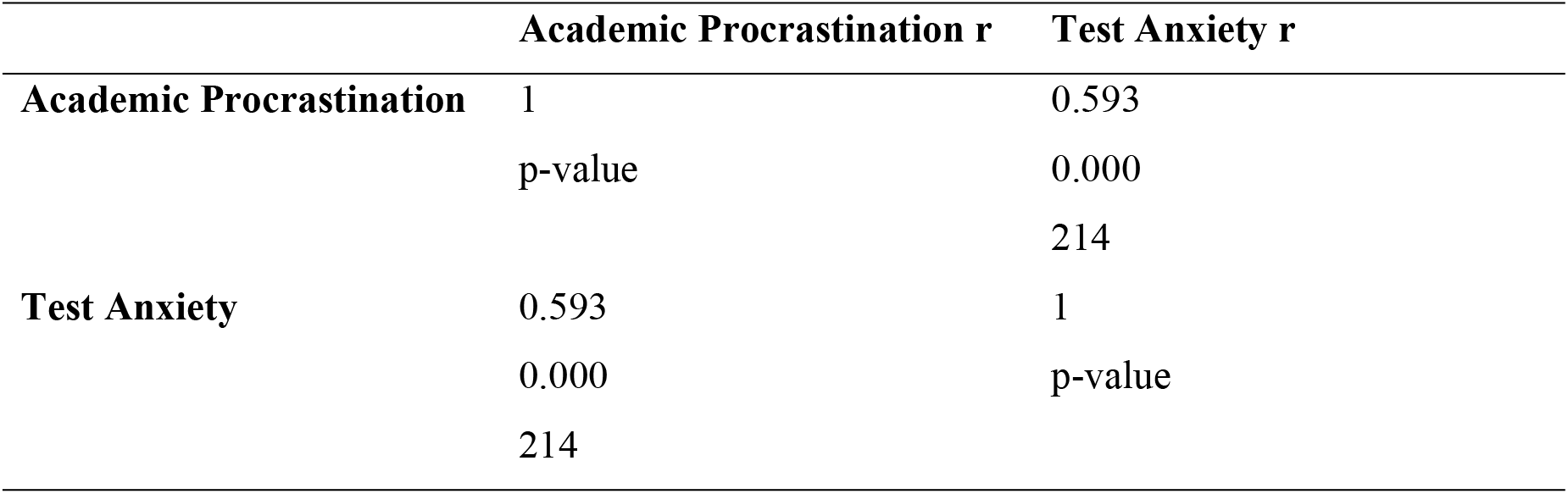
Correlation between Procrastination and Test Anxiety.

#### Binary and Multiple Logistic Regression of variables associated with Test Anxiety

Table below represents the simple binary and multiple logistic regression of variables with Test Anxiety. Those variables that had the significant association with Test Anxiety were further analyzed and Crude odds ratio and Adjusted odds ratio were calculated. The odds of having Test Anxiety among respondents whose maternal education level was secondary/more were 1.040 times (95% CI 0.526-2.056 p-value=0.910) more likely than maternal education level less than secondary. Similarly, the odds of having Test Anxiety among respondents whose paternal education level was secondary/more were 1.071 times (95% CI 0.463-2.476 p-value=0.873) more likely than paternal education level less than secondary.

#### Correlation between Academic Procrastination and Test Anxiety

From the table 18 it shows the moderate degree of positive correlation between the Academic Procrastination and the Test Anxiety (r=0.593) where students with test anxiety is greatly affected by the academic procrastination.

## Discussion

This chapter presents the results of a study that aimed to assess procrastination and test anxiety and its associated factors among medical students (MBBS) in CMC, Bharatpur. This research finding is organized according to the study’s five objectives, which focused on exploring the procrastination and examine the levels of test anxiety experienced by medical (MBBS) students of medical colleges, identifying their associated factors and analyzing the relationship between procrastination and test anxiety among medical (MBBS) students. The results are compared with previous studies that support or contrast with the current findings.

### Procrastination and its associated factors

In this study, the findings reveal that those who are staying on their own or with flat mates have higher mean score (75.37) compared to living at home with family (74.09) and hostel (67.98) with the significance of **p-value=0.001**. The previous study conducted in India observed the effects of living status on academic procrastination the significance is **p=0.135**. Since p=0.135> p=0.05,(29) which states there is not any difference between the levels of academic procrastination among people living with family and living away from home. It shows the **differences in association of residence and procrastination** between two studies. As nearly two third of the respondents from relevant study were living with family while less than one sixth of the respondents from present study were living with family which might be the possible reason.

In this study, among **sociodemographic factor** neither of the variable showed significant association with procrastination likewise compared to the mean **male had higher procrastination than that of female** (mean **±** SD; male = 72.35 **±** 13.949, female= 70.05 ± 14.884) and past study in Iran showed **similar result** with (mean **±** SD; male= 81.05 **±** 14.00, female= 79.58 ± 10.43).(30) The observed higher levels of procrastination among male students could be influenced by societal pressures that discourage seeking help or admitting difficulty. Interventions targeting procrastination should address these gender-related expectations, creating an environment where seeking support is normalized for all students, thereby improving academic outcomes.

### Test Anxiety and its associated factors

In this study, the finding shows that almost two fifth of the respondents whose **parental education level** (maternal and paternal) secondary/more had test anxiety with significance p-value=0.036 and 0.048 respectively. One of the previous studies in Iran showed that **families** in which at least one parent has an **academic degree**, were **perceived test anxiety** by students to be more involved compared to non-academic families.(31) The **similarity between** the studies where both the findings reflect greater awareness of more educated parents to the importance of higher education, to a point of **placing counterproductive pressure** on their children. Similarly, the previous study in Ajman, UAE shows that among the students with family pressure almost eight out of ten students were having test anxiety (79.7%).(32) This shows the level of impact family play in their children’s academic decisions which pressurize them to leading to worry about their academic evaluation.

As one of the previous studies in Caribbean Netherland, shows that there was significant association between gender and high-test anxiety (p < 0.001), with a higher prevalence among females (47.9%) compared to males (22.5%). Although non-significant, its prevalence was higher among the 20 years old (34%). Anxiety decreased as the students progressed to higher years of studies (37.9% in the third year to only 9.1% in sixth year, p=0.073).(33) However in present study comparing sex; male (40.4%) had higher percentage in test anxiety than female (37.2%), age wise; students aged less than 20 years had test anxiety percentage (44.6%) and according to year of education not a continuous pattern as it shows higher percentage in first year students (46.3%) and fourth year students (46.2%) than that of second year students (33.9%) and fifth year (30.8%). Previous research showed higher anxiety among females and younger students, with decreasing anxiety as students progressed in their studies. However, the current study found slightly higher anxiety among males, highest anxiety among younger students, and inconsistent patterns across academic years. The implications of these findings could include the need for specific interventions to address test anxiety among students, considering variations based on gender, age, and academic year.

### Procrastination and Test Anxiety

In this study the it shows the strong correlation between the Academic Procrastination and the Test Anxiety with the positive correlation value (r=0.593) **moderate correlation** value which illustrates that students with test anxiety are greatly affected by the academic procrastination. The past study in Iran where was a statistically **significant, positive correlation** between test anxiety score and procrastination score (*P* value = 0.000, Pearson’s correlation co-efficient = 0.443).(31)

Both the study concluded that procrastination leads to test anxiety because of the past **negative evaluative experiences** and lack of confidence of test anxious students. The students may also believe that their efforts may not yield the desired results, which may lead to last minute studying to pass the examinations, further leading to anxiety. Additionally, same as the present study, the study in Lebanon, supports correlation between procrastination and test anxiety is (r=0.341) moderate and positive, indicating that students who procrastinate more tend to experience higher levels of test anxiety.(34) This supports the hypothesis and aligns with previous research. Understanding these mechanisms can help educators and students address procrastination behaviors and reduce test anxiety.

#### Limitation of the study

In the standard tool, The Academic Procrastination Scale there are altogether six domains but those aren’t elaborated so it was limited to total procrastination as an output.

## Conclusion

This institution based cross-sectional study gives valuable information on the procrastination and test anxiety and its associated factors among medical students (MBBS) of Chitwan Medical College using semi-structured questionnaire. A total of 214 participants were included in the study using probability proportionate sampling technique. Permission was obtained from the principal of CMC and Institutional Review Board of MMIHS IRC. Data was collected using self-administered questionnaire, imported to SPSS, and analyzed using appropriate statistical techniques.

The study revealed that a higher percentage of respondents’ mothers were homemakers, with very few involved in foreign employment, whereas a higher percentage of fathers were involved in the business sector, with very few being homemakers. Additionally, the study showed that almost half of the respondents were staying at hostels, followed by less than one third living close to campus on their own or with flat mates, and less than one fifth were living at home with their family. Furthermore, the study found that males exhibited higher levels of procrastination and high-test anxiety than females, with variations observed across different ethnicities, with Muslims and Dalits tending to have higher levels of procrastination and high-test anxiety despite being a minority among respondents. Regarding age, an increase in age was associated with increased procrastination and decreased test anxiety. The study also indicated that the higher the parental education level, the higher the level of procrastination among students.

In conclusion, almost two-fifths of the students were experiencing test anxiety. Procrastination exhibits significant differences across factors such as student residence, alcohol use, and smoking/tobacco product use, indicating that individual factors contribute more to academic procrastination than socio-demographic factors. Test anxiety also demonstrates significant differences across factors such as maternal and paternal education levels; with students whose parents have secondary or higher education levels showing higher test anxiety. The relationship between procrastination and test anxiety reveals a moderate correlation, with academic procrastination and test anxiety having a positive correlation value (r=0.593), illustrating those students who procrastinate more tends to have higher test anxiety.

## Data Availability

All relevant data are within the file RashmiKandel_DATASET.

## Acknowledgement

Pleased to have an opportunity to conduct the study on “Procrastination and Test Anxiety among Medical Students of Medical Colleges”. The completion of this research could not have been possible without the supervision, guidance and participation of all people. This report is the joint outcome of all individuals and thus everyone deserves to be acknowledged. Immense Gratitude towards Chitwan Medical College, Bharatpur; Chitwan for granting permission to conduct research among Medical Students (MBBS) of Chitwan Medical College, for granting permission of all respondents’ involvement in the data collection.

In this regard, Manmohan Memorial Institute of Health Sciences, the entire administration team, Campus Chief Prof. Mr. Dharma Khanal, Department of Public Health, Associate Prof. Khadka Prasad Shrestha (Head of Department), Coordinator Lecturer Laxmi Gautam was duly acknowledged for providing with the opportunity to carry out the study and get acquainted with research process.

Sincere gratitude towards the supervisor, Professor Dr. Sujan Babu Marahatta for his constant guidance, valuable suggestions, constructive feedback and word of encouragement throughout the research. Moreover, special thanks to Lect. Sudip Khanal for his valuable suggestions for deriving the output of the research and also all the faculty members for valuable comments, guidance and encouragement.

Individuals who generously gave their time and shared their knowledge to help complete this research are also acknowledged. The insights and feedback from family and colleagues were essential in shaping the direction and scope of the project. It was an honor and privilege to work with such a remarkable team.

Rashmi Kandel

## Notes

### Competing Interest Statement

The authors have declared no competing interest.

### Funding Statement

The author(s) received no specific funding for this work.

### Author Declarations

This study received ethical approval from the Institutional Review Committee of Manmohan Memorial Institute of Health Sciences, Kathmandu, Nepal (Approval Number: NEHCO/IRC/080/051). Written informed consent was obtained from all participants before their participation in the study. All procedures performed were in accordance with relevant institutional and national research ethics guidelines.

